# Pain state-dependent multimodal assessment in non-specific low back pain: a pseudorandomized study design with implementation insights

**DOI:** 10.64898/2026.08.26.26359760

**Authors:** Beatriz Chozas Barrientos, Madeleine Hau, Laura Sirucek, Anke Langenfeld, Martina Wehrli, Brigitte Wirth, Niklaus Zölch, Jan Devan, Stefan Dudli, Petra Schweinhardt

## Abstract

**Background:** Fluctuations in pain intensity are intrinsic to non-specific chronic low back pain (nsCLBP). Nevertheless, pain fluctuations have rarely been considered when investigating pathophysiological mechanisms. Therefore, a novel study protocol was developed and implemented to systematically assess the impact of fluctuating pain states on pain-related measures.

**Methods:** The final study cohort consisted of 45 nsCLBP patients and 47 age- and sex-matched healthy controls (HCs). Patients participated in three visits, conducted during different pain states (i.e. clinically relevant pain, low-intensity clinical pain / pain-free, clinically irrelevant pain induced using a Qutenza 8% capsaicin patch). Pain fluctuations were monitored through online assessments every four days and guided the pseudorandomized visit scheduling. HCs participated in a single visit. Each study visit comprised a multimodal battery of pain-related measures.

**Results:** 93.33% of patients completed all three visit types in a pseudorandomized order (χ²=1.50, p=0.826). Visit scheduling was possible due to the high self-report adherence (median=93.48%), unrelated to self-report burden (ρ=-0.097, p=0.53). Study visits were conducted during different pain states, as indicated by: i) the significantly higher low back pain intensity in the clinically relevant pain visit (mean[SD]: 3.98[0.90]), compared to the low-intensity clinical pain (1.03[0.86]) and clinically irrelevant pain (1.13[0.82]) visits (p-values<0.001), as well as by ii) the successful induction of a moderate-to-high clinically irrelevant pain across assessments.

**Conclusion:** Despite scheduling complexity and pain state transition uncertainty, a pain state-dependent pseudorandomized study design is feasible and could improve the understanding of nsCLBP mechanisms.

## 1. Background

Low back pain is highly prevalent worldwide and is associated with substantial disability [15]. Approximately 90% of cases are categorized as non-specific chronic low back pain (nsCLBP) [12], indicating the absence of a clearly identifiable underlying cause or pathology, although degenerative or other structural changes may be present. In this context, research has increasingly focused over the past decades on mechanistic explanations within a biopsychosocial framework [19], aiming to better understand what drives and maintains nsCLBP and to develop more targeted treatment approaches. In fact, compared to healthy controls (HCs), numerous alterations associated with nsCLBP have been found regarding psychological state [11], psychophysical measures of pain sensitivity and endogenous pain modulatory mechanisms [4,41], brain structure and function [32] as well as neurochemical characteristics [69] and blood markers [42]. Furthermore, recent large dataset studies suggest that some of these factors are relevant for chronic pain prognosis [17,58]. However, the pathophysiological relevance in the development and maintenance of nsCLBP remains unclear.

This uncertainty is partly driven by the inherent complexity and dynamic nature of pain, which arises from the interaction of multiple peripheral and central processes that may fluctuate over time and across individuals [21], leading to corresponding fluctuations in the pain experience itself [47]. However, the influence of ongoing clinical pain on experimental pain-related measures remains largely understudied in human research.

This may be because frequent pain fluctuations in musculoskeletal conditions [51] make the identification of and the recruitment during distinct clinical pain states difficult, thereby complicating the systematic investigation of state-dependent effects on pain-related measures. Some studies have attempted to tackle this issue by experimentally exacerbating clinical pain and examining pain-related measures across different pain levels [14,37]. However, few studies have investigated pain-related measures during naturally occurring fluctuations in clinical pain or across distinct clinical pain states. In fact, to the best of our knowledge, only two studies have addressed this gap in chronic pain patients with musculoskeletal conditions. These studies showed that in nsCLBP [40] and fibromyalgia [8] patients, differences from HCs were mainly observed during ongoing clinical pain and not during pain-free states. This suggests that the current pain state critically influences experimental pain-related measures. However, the interpretation of these findings is constrained by methodological limitations, such as fixed visit order and between-subject comparisons. Furthermore, these studies do not provide insights into whether the observed alterations reflect (1) trait-like pathophysiological mechanisms independent of a clinical pain episode, (2) transient pathophysiological state-dependent changes linked to clinical pain episodes, or (3) more general effects due to the sheer presence of pain that are unrelated to pathophysiological meaningful states.

To address this, a study protocol was developed to assess multiple pain-related measures in nsCLBP patients across distinct pain states, and in a group of HCs, with the aim of improving understanding of pathophysiologically relevant mechanisms underlying nsCLBP (Figure 1). The primary aim of the current manuscript is to describe the rationale and methodological framework of the study design, and to summarize key operational and feasibility characteristics observed during its implementation.

**Figure 1.**
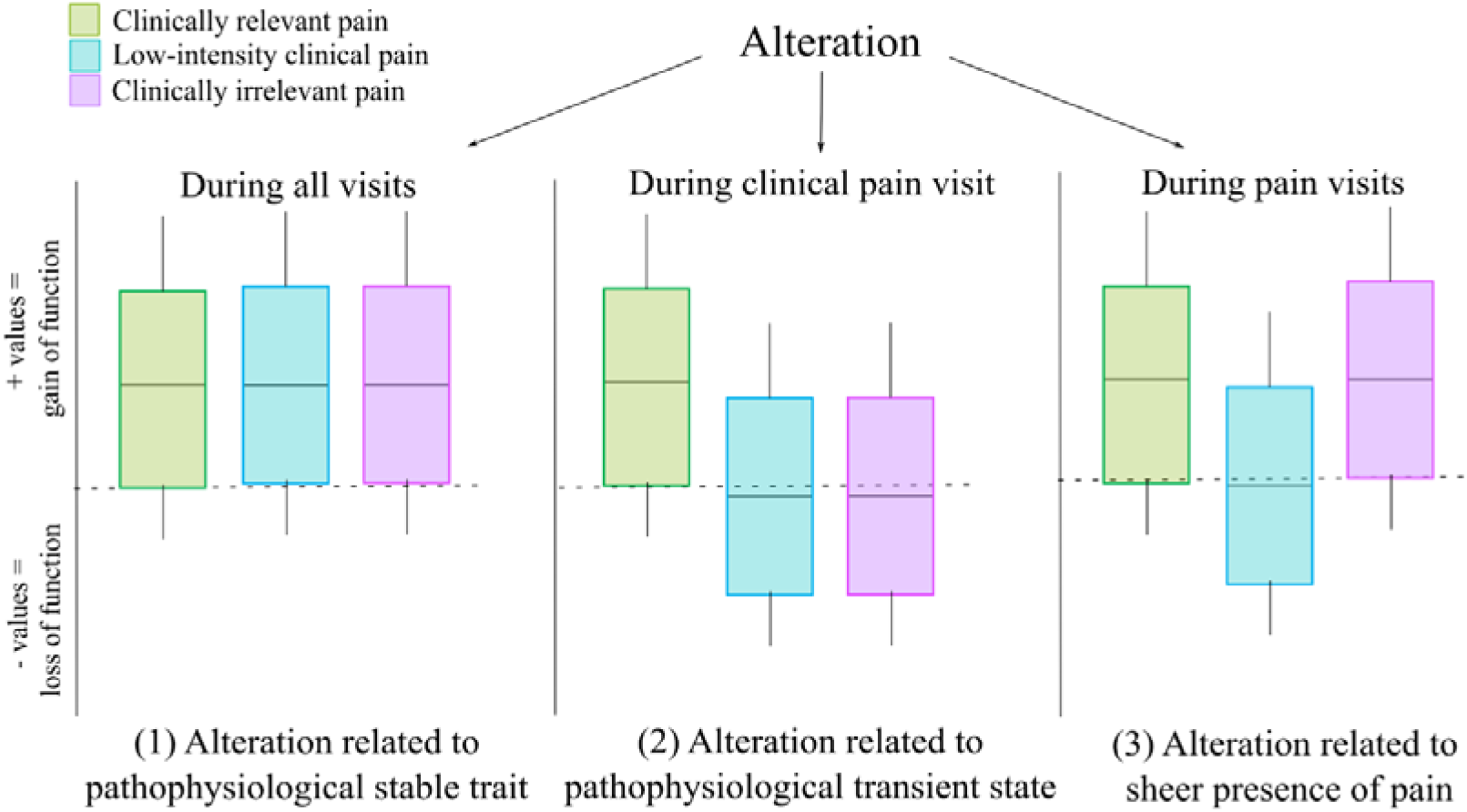
Conceptual model for disentangling the meaning of observed alterations based on the study visit in which they occur. The dashed horizontal line represents the mean value of the healthy control reference cohort. Colored boxplots represent the distribution of patient values at each visit type.

## 2. Methods

The current study implemented a pseudorandomized repeated-measures design in nsCLBP patients. Following recommendations [13], a multimodal set of pain-related measures was obtained, including clinical characteristics, psychophysical, neurophysiological, magnetic resonance imaging (MRI) and magnetic resonance spectroscopy (MRS), blood sampling and questionnaire readouts, to investigate mechanisms across different pain states. The selection of measures in the current study was informed by findings from the first study period (CRPP Pain: https://www.crpp-pain.uzh.ch/en.html). Therefore, parts of the Methods section may be similar to previously published studies within the CRPP Pain framework because similar or identical methodologies were used.

The study was approved by the local ethics committee Kantonale Ethikkommission Zürich (Nr.: 2023-00560), registered on clinicaltrials.gov (NCT06412484), and performed in accordance with the guidelines of the Declaration of Helsinki.

### 2.1 Participant Recruitment

Participants with episodic nsCLBP were recruited via Balgrist University Hospital and advertisements in Swiss chiropractic practices or UZH Marktplatz (University of Zurich online platform) between November 2023 and January 2026. Inclusion criteria for patients were age between 18 and 70 years and nsCLBP lasting for more than 3 months, without signs of specific causes for their back pain (including sensory or motor loss potentially associated with radiculopathy, infection, malignancy, rheumatic and systematic inflammatory disorders, fracture), and with an episodic time course characterized by pain-free or low-intensity pain periods, i.e. < 3 on a 0-10 numerical rating scale (NRS), and pain episodes, i.e. ≥ 3 NRS [47], lasting for several days. Age- and sex-matched HCs were recruited via UZH Marktplatz, advertisements in Balgrist Campus, and from internal participant pool registries. HCs were eligible for the study if they were pain-free or had not had low back pain for more than three consecutive days in the past year.

Exclusion criteria were any major medical or psychiatric condition that affects physical capacity or pain sensitivity (i.e., severe heart disease, diabetes, etc.) and pregnancy. The experimenter checked inclusion and exclusion criteria via phone screening. If all participation criteria were met and participants gave oral informed consent to participate, they were emailed an electronic baseline REDCap [25] questionnaire on demographics and general health (Supplementary Material 1), which had to be filled out before on-site participation.

At the beginning of the first study visit on-site, participants gave written informed consent to participate. Subsequently, a clinician (physiotherapist or chiropractor) repeated the phone screening questions and performed a bedside assessment of sensory function (vibration, thermal, pinprick, and light touch) and muscle strength in all participants. In case of any abnormal finding, the participant (patients and HCs) was excluded.

### 2.2 Study Design

The study design (Figure 2) aimed to gain insight into the impact of distinct pain states on a multimodal set of pain-related measures that can capture nociceptive processing alterations in nsCLBP. For this, participants were tested in three visits during different pain states, while HCs were tested in a single visit.

**Figure 2.**
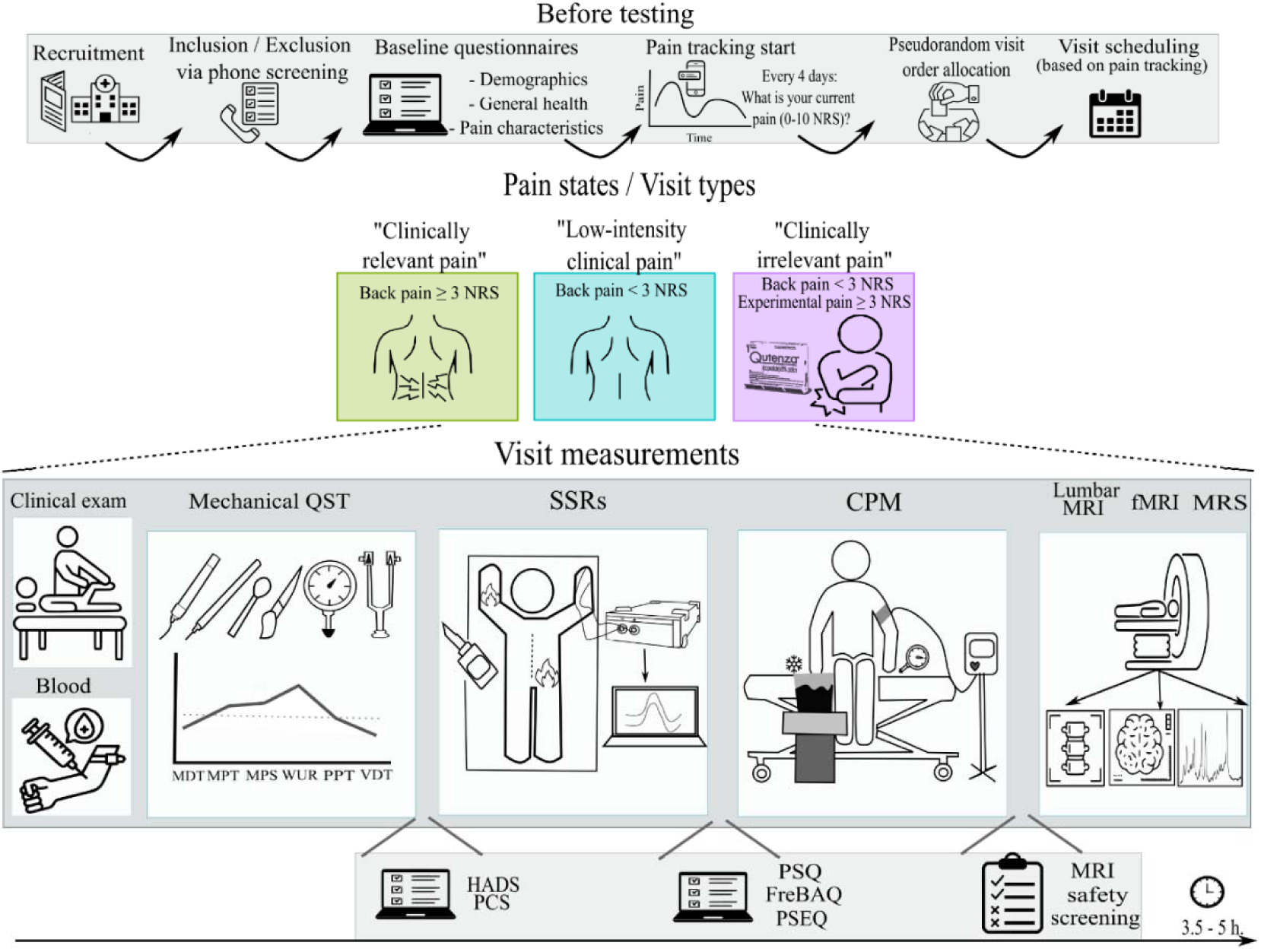
Study design. CPM: conditioned pain modulation, FreBAQ: fremantle back awareness questionnaire, HADS: hospital anxiety and depression scale, MDT: mechanical detection thresholds, MPT: mechanical pain thresholds, MPS: mechanical pain sensitivity, (f)MRI: (functional) magnetic resonance imaging, MRS: magnetic resonance spectroscopy, NRS: numerical rating scale, PCS: pain catastrophizing scale, PPT: pressure pain thresholds, PSEQ: pain self-efficacy questionnaire, PSQ: pain sensitivity questionnaire, SSRs: sympathetic skin responses, VDT: vibration detection thresholds, QST: quantitative sensory testing, WUR: wind-up ratio

One visit took place during an ongoing pain episode of at least moderate intensity, with a cutoff set at ≥ 3 NRS, conceptualized as the clinically relevant pain visit. The ≥ 3 NRS threshold was selected to identify moderate-intensity pain episodes [7], based on the Delphi consensus definition of pain episodes. One visit took place on a low-intensity clinical pain / pain-free day, with a cutoff set at < 3 NRS, conceptualized as the low-intensity clinical pain visit. Another visit took place on a low-intensity clinical pain / pain-free day, during which an experimental pain state was induced. The rationale for including such a visit in the study design was to control for confounding effects due to the sheer presence of pain. This visit was conceptualized as the clinically irrelevant pain visit. A 4x5 cm Qutenza 8% capsaicin patch (Grünenthal Pharma AG, Switzerland) applied at the dominant volar forearm was chosen as an experimental model because it allowed to induce a prolonged clinically irrelevant pain state that was present at rest but as different as possible to the patients’ clinically relevant pain, i.e., in terms of i) location: arm vs. back, ii) affected anatomical structure: skin vs. musculoskeletal tissue, iii) sensation: superficial burning-like pain vs. deep back pain.

The visit type (i.e., clinically relevant pain, low-intensity clinical pain and clinically irrelevant pain) order was pseudorandomized. nsCLBP patients were randomly assigned to one of six predefined visit sequences (Table 1), keeping a balanced number of participants assigned to each sequence. The interval between visits was adapted to the patient’s individual pain fluctuation pattern and availability.

**Table 1.** Possible visit orders randomly assigned to patients at enrollment. CR: clinically relevant, CI: clinically irrelevant, LI: low-intensity clinical pain.

| Visit order | 1<sup>st</sup> visit | 2<sup>nd</sup> visit | 3<sup>rd</sup> visit |
| --- | --- | --- | --- |
| **A** | LI | CR | CI |
| **B** | LI | CI | CR |
| **C** | CI | LI | CR |
| **D** | CI | CR | LI |
| **E** | CR | LI | CI |
| **F** | CR | CI | LI |

Fluctuations in pain intensity were tracked using an automatically-triggered REDCap email link sent to patients every four days until completion of the study with the following question: "*How severe is your current pain on a scale of 0 to 10, with 0 being no pain and 10 being the worst pain that can be tolerated?*". The experimenter consistently monitored the patients’ self-reports and scheduled each visit only when the pain state matched the one required by the pre-assigned visit order. In addition, to maximize scheduling success, participants were instructed to proactively contact the experimenter whenever a clear pain state fluctuation occurred. These communication pathways were intentionally kept flexible (i.e., phone call, SMS, email, messaging applications), prioritizing rapid pain state capturing over rigid communication standardization.

### 2.3 Pain-related Measures

The pain-related measures are presented in the chronological order in which they were conducted within a visit, unless specified otherwise. The chronological sequence was chosen a priori to account for potential interactions between assessments and ensure consistent experimental conditions across visits. Clinical characteristics served to characterize the cohort and were therefore only assessed once, while all the other readouts were assessed at every visit, unless specified otherwise, with the aim to disentangle whether alterations observed in patients in comparison to HCs reflect state-related or trait-related mechanisms.

#### Test Areas and Clinical Characteristics

Clinicians anatomically identified the patients’ most painful area in the lower back (MP) and a rostrally adjacent pain-free area as close as possible to MP (AD). The location of these areas later served as test sites and was measured as the distance (in cm) from the posterior superior iliac spine on the x- (left-right) and y- (cranial-caudal) axes. This allowed to identify the same test sites in subsequent patient visits and to reproduce the test sites in the HC individually matched to each patient. A remote pain-free area, i.e., non-dominant hand or volar forearm, depending on the pain-related measure, served as the control (CON) area.

During patients’ clinically relevant pain visit, a clinician performed a comprehensive clinical examination developed following an evidence-based diagnostic classification framework [63]. This clinical examination included provocation tests and neurological and functional assessments, with the goal to identify the patients’ most likely nociceptive source. Patients were asked to draw their typical spatial pain extent on a body chart (frontal and dorsal views) [54]. On a separate body chart, patients drew their momentary pain extent at the beginning of each visit.

Additional clinical pain characteristics were collected via the online baseline questionnaire in REDCap (Supplementary Material 1), which included general information on pain intensity, beliefs about pain origin, and worries associated with pain, as well as specific questionnaires: painDETECT [18], inflammatory pain pattern screening [3], Widespread Pain Index (WPI) [67] and Central Sensitization Inventory (CSI) [38].

To further characterize the cohort, all participants underwent a standard clinical lumbar MRI, performed before or after the functional brain MRI depending on scanner availability, in a 3T Siemens scanner lasting approximately 15 minutes and consisting of sequences measuring sagittal T1, sagittal T2, sagittal T2 with fat suppression (DIXON technique, [35]), and axial T2.

#### Blood Sampling

Whole blood was collected by a study nurse into three tubes: BD PAXgene Blood RNA (2.5 mL), serum separating (6 mL), and K2-EDTA BD (4 mL) for analyses of whole blood transcriptomics, serum proteins, and cellular composition, respectively [16]. Blood was sampled during patients’ clinically relevant pain and low-intensity clinical pain visits, but not in the clinically irrelevant pain visit, as potential changes in blood marker levels induced by the Qutenza 8% capsaicin patch were not expected to happen within the visit’s time frame. Blood was processed according to the biospecimen collection protocol of the NIH Low Back Pain Consortia (BACPAC) [16].

#### Quantitative Sensory Testing (QST)

A shortened version of the original German Research Network on Neuropathic Pain (DFNS) battery [53] using mechanical stimuli was implemented. Mechanical stimuli are the most widely used modality in musculoskeletal research [4], as they can activate nociceptors in different relevant tissues, including deep structures, which are typically involved in conditions like nsCLBP. The QST battery consisted of mechanical detection thresholds (MDT), mechanical pain thresholds (MPT), mechanical pain sensitivity (MPS), dynamic mechanical allodynia (DMA), wind-up ratio (WUR), pressure pain thresholds (PPT) and vibration detection thresholds (VDT). Due to time limits within the larger study protocol, MPS and DMA, as well as WUR, were evaluated with 3 (instead of 5) blocks. Pain intensity was rated on an NRS from 0 “no pain” to 100 “the most intense pain sensation imaginable”, consistent with the DFNS QST protocol. This 0-100 NRS was used across all experimental pain-related measures to avoid switching between scales, while the 0-10 NRS was used for clinically oriented pain ratings (e.g., low back pain), as it is more commonly used in clinical practice and familiar to patients.

Mechanical QST was conducted at three body areas. The nondominant hand was the CON area and was always tested first, as per DFNS recommendations. This was followed by the MP and the AD areas. The test order of MP and AD areas was pseudorandomized and kept constant for each visit of a given patient and for the HC individually matched to this patient. Assessing mechanical loss and gain of function at these three body sites allows to potentially disentangle peripheral vs. central, including spinal or supraspinal, mechanisms that might be driving observed sensory alterations [57].

### Sympathetic Skin Responses (SSRs)

SSRs were triggered using noxious contact heat stimuli applied with a 27 mm diameter CHEPS thermode (Pathway Pain and Sensory Evaluation System®, Medoc Advanced Medical Systems, Ramat Yishai, Israel). The thermode’s baseline temperature was set to 42°C and the destination temperature was 52°C (70°C/s ramp). If participants could not tolerate the baseline stimulation, a 35°C baseline temperature was used. The thermode was held manually and slightly moved between stimuli to minimize peripheral adaptation or sensitization. Participants verbally rated the painfulness of each contact heat stimulus on an NRS 0-100 scale. SSRs were recorded in the prone position using a pair of 8 mm Ag-AgCl electrodes with Ten20 conductive paste (ADInstruments®, Dunedin, New Zealand) fixated on the first phalanges of the index and ring fingers of the dominant hand using medical tape (3M™ Transpore™). SSRs were measured as the voltage difference (µV) between the active and the reference electrodes, which were connected to a PowerLab device (ADInstruments®, Dunedin, New Zealand), and time-locked to the contact heat stimuli.

SSRs were recorded for contact heat stimulation at two testing sites: at MP and at the volar forearm of the non-dominant arm (CON). At each testing site, 15 contact heat stimuli were applied with an 8 – 19 s interstimulus interval. There was a 2-minute break between testing sites, and the test order of MP and CON areas was pseudorandomized and kept constant for each visit of a given patient and for the HC individually matched to this patient. Assessing SSRs, a neurophysiological measure of sympathetic autonomic activity [62], at these two body sites allows the investigation of potential arousal- and autonomic-related differences in nociceptive processing between a clinically affected region and a remote, non-painful control site.

### Conditioned Pain Modulation (CPM)

CPM paradigms investigate the change of pain perception, defined as the CPM effect, of a phasic noxious test stimulus at baseline compared to during or after the application of a heterotopic tonic noxious conditioning stimulus [52].

Pressure test stimuli were applied on the thenar eminence of the non-dominant hand using a Force Dial FDK 10 algometer (Wagner Instruments) with a custom-made spherical tip of 1 cm diameter. At baseline, pressure pain thresholds (PPTs) were determined three times by using a continuous stimulus increasing at a rate of 0.5 kg/s [53]. Based on the arithmetic mean of baseline PPTs and following a procedure based on internal pilot testing, a suprathreshold stimulus intensity that aimed to elicit a pain rating of 40 (35 – 45) on an NRS 0-100 scale was determined using a maximum of four stimulations to minimize sensitization. Here, the pressure stimulus was applied for 1s. Next, blood pressure, which has been shown to be associated with pain tolerance and pain inhibition [9] was measured on the non-dominant arm using a Mindray VS-800 device. After this measurement, one suprathreshold pressure stimulus at the predetermined intensity was applied and rated. The three readouts performed at baseline (PPT, suprathreshold pain rating and blood pressure) were measured again during and after a water bath that was either cold (9°C ± 0.5), serving as the conditioning stimulus, or lukewarm (32°C ± 0.5), serving as a control condition. There was a 10-minute break between the two water baths. The order of cold and lukewarm water baths was pseudorandomized and kept constant for each visit of a given patient and for the HC individually matched to this patient. Each water bath allowed to calculate parallel (during – baseline) and sequential (after – baseline) CPM effects of all three readouts.

The CPM effect is considered to reflect endogenous descending pain modulation in humans [48]. Assessing pain modulation in a non-affected region had the aim to determine whether the efficiency of descending pain modulatory pathways, which might be altered in chronic pain [49], is compromised beyond the site of pain, minimizing the influence of local nociceptive processes.

### Psychological and Sensory Questionnaires

Patients electronically completed the following questionnaires assessing psychological constructs or sensory aspects related to the experience of pain: Hospital Anxiety and Depression Scale [70] (HADS), Pain Catastrophizing Scale [10] (PCS), Pain Sensitivity Questionnaire [55] (PSQ), Fremantle Back Awareness Questionnaire [64] (FreBAQ), and the Pain Self Efficacy Questionnaire [46] (PSEQ). HCs completed all questionnaires except the PSEQ because it assesses functioning despite chronic pain, which is not applicable to this group.

### Functional Magnetic Resonance Imaging (fMRI)

The fMRI session, conducted on a 7T Siemens MAGNETOM scanner, included a structural scan using a 7 min T1-weighted magnetization prepared 2 rapid gradient echo sequence (MP2RAGE; TR: 6000 ms, TE: 1.99 ms, voxel size: 0.7 mm isotropic, field of view (FOV): 240 mm), a field map (gradient echo sequence, TR: 4.5 ms, TE: 1.02 ms / 3.06 ms, voxel size: 4 mm isotropic, FOV: 256 mm) and a 14 min whole-brain resting-state fMRI using a T2*- weighted echo-planar sequence (repetition time (TR): 955 ms, echo time (TE): 22 ms, voxel size: 1.6 mm isotropic, FOV: 208 mm, flip angle: 45°, 900 volumes, multiband acceleration factor: 5). Due to artifacts that were detected in the functional data of the first subjects, a fat saturation was added after the first six sessions. The total scanning session lasted approximately 25 min. Participants were instructed to lay still in the scanner with eyes open looking at a fixation cross. Structural and functional MRI data will be preprocessed and analyzed using FSL version 6.0.7 [29].

Brain morphology and resting-state functional connectivity have been extensively used to assess supraspinal involvement in chronic pain [33]. Assessing these measures across different pain states may provide further insight into the neural mechanisms underlying chronic pain and help characterise how pain perception and its variability relate to brain structural [2,59] and functional [30,31,37,68] measures.

### Magnetic Resonance Spectroscopy (MRS)

The MRS session, conducted on a 3T Philips Achieva scanner, included a structural scan using a 7 min T1-weighted magnetization prepared rapid gradient echo sequence (MPRAGE; TR: 8.1 ms, TE: 3.7 ms, voxel size: 1 mm3 isotropic, FOV: 240x160x240 mm3 (APxLRxFH), flip angle: 8°, shot interval: 3000 ms) that preceded the MRS acquisition and was used for placement of the MRS voxel (8.8 × 10.2 × 12.2 mm³ (1.1 mL)). The voxel was placed by the same examiner at the periaqueductal gray (PAG) based on anatomical landmarks. Five pairs of outer-volume suppression pulses and voxel-specific flip-angle calibration were applied to improve localization and optimize signal-to-noise ratio. Water suppression was performed using VAPOR, and second-order automatic pencil-beam shimming was applied prior to MRS acquisition. A single-voxel proton MRS (^1H-MRS) acquisition from the PAG lasting 24 mins was conducted using a standardized semi-localization by adiabatic selective refocusing (sLASER) sequence (TR: 2500 ms, TE: 30 ms, spectral width: 2000 Hz, and 2048 spectral points). For each participant, 512 water-suppressed averages were acquired in eight blocks of 64 averages. At the beginning of each block, an unsuppressed water reference scan was acquired, resulting in eight short-TR water reference measurements. Compared to standard point-resolved spectroscopy (PRESS), sLASER is less sensitive to field inhomogeneities and exhibits substantially reduced chemical shift displacement artefacts [50]. Additionally, the shorter effective T2 relaxation times and improved localization may facilitate the detection of J-coupled metabolites such as Glutamate and Glutamine [24]. Following the MRS acquisition, two additional unsuppressed long-TR water reference scans (TR: 10 and TR: 12 s) were acquired from the same voxel. Head motion was monitored using a markerless motion-tracking system (TracInnovations, Ballerup, Denmark). The total scanning session lasted approximately 40 min. Participants were instructed to lay still and could listen to music on the radio to increase comfort in the scanner. Spectroscopic data will be preprocessed using ReconFrame (GyroTools LLC, Zurich, Switzerland).

The PAG is a brainstem region central to endogenous pain modulation [5]. The aim was to measure metabolite concentrations in the PAG, with a particular focus on glutamate and GABA, which are primary mediators of the brain’s excitatory/inhibitory balance. This excitatory/inhibitory balance may be altered in chronic pain, as suggested by animal [34] and human [56] studies.

### 2.4 Statistical Analyses

Statistical analyses were carried out using R statistical software (version 4.4.1 for Windows).

### Study Implementation Insights

To characterize the feasibility and burden of repeated self-reporting, the following metrics were calculated for each patient. (i) *Number of expected reports* = *duration of participation (days) / 4 (days; frequency of self-reporting)*. The number of *expected self-reports* is also referred to as *self-report burden*. For example, a patient completing the study in 80 days would have 80 / 4 = 20 expected self-reports, which is a higher self-report burden than a patient completing the study in 40 days, for whom 40 / 4 = 10 self-reports would be expected. (ii) *Self-report adherence (%)* = (*N successfully completed self-reports [+ 1 day delay]* / *N expected self-reports) * 100*.

To characterize the temporal dynamics of pain fluctuations, the pain state stability of each patient was assessed by calculating the following metrics. (i) *Number of transitions between pain states*, characterized as a change from moderate pain intensity ≥ 3 NRS to a low-intensity clinical pain < 3 NRS, or vice versa, during study participation. (ii) *Mean duration of self-reported states (in days)*, where the duration of each consecutive period spent in the same pain state (i.e., low-intensity clinical pain either < 3 NRS or clinically relevant pain > 3 NRS) was calculated for each patient as: *date last pain rating in a certain state – date first pain rating in a certain state*. For patients with multiple pain state transitions, the durations of each consecutive period spent in the same pain state were averaged to obtain an individual mean pain state duration.

At the group level, descriptive summary statistics appropriate to the data distribution were conducted.

### Plan of Study Pain-Related Measures

Statistical analyses will be chosen according to data distribution, as assessed using histograms and quantile–quantile plots. Changes in pain-related measures across different pain states (clinically relevant pain vs. low-intensity clinical pain vs. clinically irrelevant pain) will be analyzed using appropriate models or tests and will be further compared in post-hoc analysis if needed. Participants will be considered as random effects and pain state as fixed effects. Visit number, age, sex or other analysis-specific variables will be included as covariates. Exploratory analysis investigating the relationship between pain-related measures and psychosocial or clinical characteristics will be performed when relevant.

## 3. Results – Protocol Implementation

The current Results section reports measurable feasibility outcomes derived from the implementation of the novel study protocol, while results of the study’s pain-related measures will be reported in subsequent manuscripts. Additionally, important operational considerations relevant to pain state-dependent research will be discussed.

### 3.1 Participant Adherence

Out of the recruited 62 nsCLBP patients and 54 HCs who orally consented to take part in the study and received the online baseline questionnaires, 11 participants never signed the informed consent and therefore were not formally enrolled due to: underestimation of time burden and visit planning incompatible with personal schedule (5 patients and 3 HCs), failure to report an exclusion criteria on phone-screening and later communication via email (1 HC), never answered the online baseline questionnaire nor the experimenter’s attempts to make contact (1 HC), and lack of stable enough pain fluctuation for visit scheduling (1 patient).

Out of the enrolled 56 nsCLBP patients and 49 HCs who signed the informed consent on-site, 8 participants were excluded from the study due to: suspected psychiatric condition (1 patient and 1 HC) or malingering (1 patient), abnormal sensory findings at the bed-side exam (1 HC), possibility of specific reasons for CLBP identified during clinical screening (4 patients: two with moderate / severe scoliosis [28], one with spondyloarthritis, and one with suspicion of radiculopathy). Finally, 5 patients participated in one study visit but discontinued study participation (1 due to time burden, 1 stopped answering to experimenter’s contact attempts, 2 were unwilling to go through MR scanning again, and 1 due to lack of pain-free periods within study duration).

The final study cohort consisted of a total of 45 nsCLBP patients (72.58% of the initially recruited patients) and 47 HCs (87.04% of the initially recruited HCs). For details on the recruitment source of the final participant sample, see Table 2.

**Table 2.** Recruitment sources of the final participant sample. Data represents participant count (%) within the respective cohort. nsCLBP: non-specific chronic low back pain, HCs: healthy controls, UZH: University of Zurich, ISR: integrative spinal research.

| Recruitment Source | nsCLBP | HCs |
| --- | --- | --- |
| Balgrist Chiropractic Clinic | 17 (37.78) | - |
| Wädenswil Chiropractic Practice | 6 (13.33) | - |
| Patient magazine | 1 (2.22) | - |
| UZH Marktplatz (online platform) | 17 (37.78) | 16 (34.04) |
| Mouth to mouth | 1 (2.22) | 13 (27.66) |
| Internal ISR / Balgrist Campus | 3 (6.67) | - |
| Participant pool registry | - | 18 (38.30) |

### 3.2 Pain State Tracking

Clinical low back pain intensity fluctuations self-reported by each patient are displayed in Figure 3. The median number of pain self-reports per patient was 17 (interquartile range [IQR]: 12–25), with the number of ratings ranging from 1 to 83. The median self-report adherence was 93.48% (IQR: 66.67 – 98), with adherence ranging from 3.74% to 100%. To explore whether self-report adherence varied as a function of self-report burden (i.e., number of expected self-reports), a Spearman correlation was computed between self-report adherence percentage and number of expected self-reports. This exploratory analysis showed no significant association (ρ = - 0.097, p = 0.53). Because self-report adherence is calculated as the number of successfully completed reports relative to the number of expected reports, some association between the two metrics could theoretically be expected. Therefore, the absence of a significant correlation further supports that self-report adherence was not substantially influenced by self-report burden.

**Figure 3.**
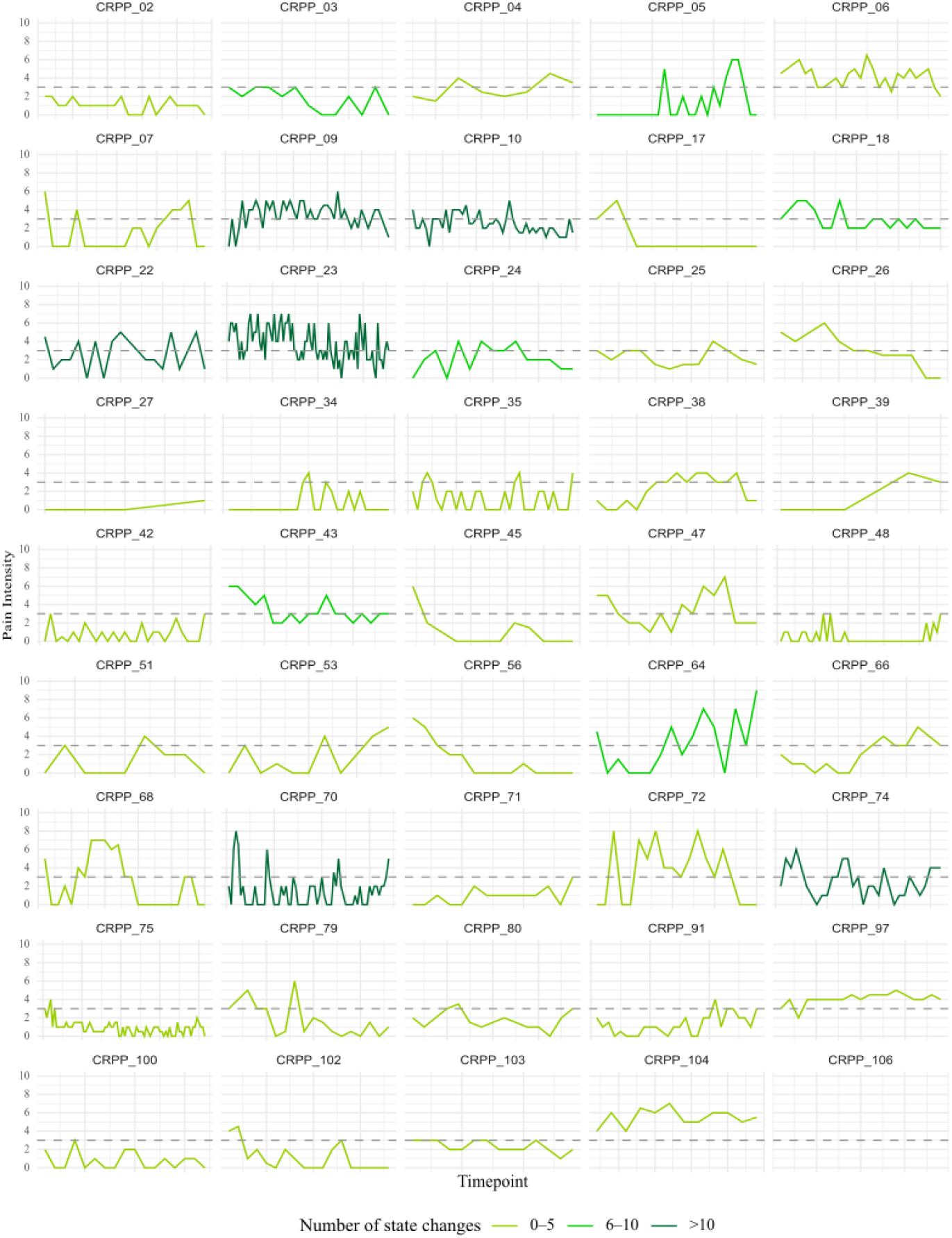
Clinical low back pain intensity fluctuations of each patient (individual plots) on a numerical rating scale from 0-10, self-reported via a REDCap email link. The dashed horizontal line (y = 3) represents the threshold delimiting a clinically relevant low back pain episode versus a low intensity / pain free period. No units are displayed in the horizontal axis, which represents time (in days), because the time frames during which patients reported their clinical pain intensity was not fixed. Indeed, it varied greatly for each patient, ranging from 8 (CRPP_27) days to 533 (CRPP_23) days. CRPP_106’s panel is empty because they provided only one pain rating using the self-report system, while a minimum of two ratings are needed to understand a pain fluctuating pattern.

Based on self-reports, patients presented a median of 4 pain state transitions (IQR: 2 – 6), with the number of transitions ranging from 0 to 32. In four patients, a pain state transition was not identified via the pain tracking system due to: real lack of pain state change (one patient was recruited from the Balgrist chiropractic clinic but never experienced a moderate low back pain episode again during the study duration, leading to an incomplete study participation with 2/3 study visits), lack of sensitivity of the automatic self-report system (two patients reported only pain intensity below or above threshold, but proactively contacted the experimenter via other channels to communicate a clear change in pain state and to promptly schedule a study visit), and insufficient self-reports on the automatic system (one patient, consistently contacted the experimenter via other channels and provided only one pain rating using the self-report system). At the group level, the median duration (days) during which patients (excluding the patient providing one single self-report) consecutively reported pain intensity ratings belonging to a same pain state was 15.75 (IQR: 7.33 – 22.30, ranging from 1 to 206).

### 3.3 Pain State Visit Scheduling

Out of the final nsCLBP cohort with 45 patients, 93.33% (n = 42) completed all three visits and 6.67% (n = 3) completed only two visits. Two of them lacked the clinically relevant pain visit due to: lack of a moderate low back episode during the study duration and impossibility of scheduling a visit during a moderate low back episode. One patient did not complete the clinically irrelevant pain visit because they entered a period of strong low back pain that interfered with daily functioning and ability to keep up with everyday responsibilities. As a result, their engagement in the study declined, leading to a discontinuation of pain self-reports. Via other communication channels, the experimenter was nevertheless informed that within the timeframe in which the study was ongoing, pain intensity had not remitted. Therefore, the low-intensity clinical pain criteria required for that visit were not met.

The distribution of visit types across visit number (Figure 4) was examined with a chi-square test of independence. Visit types (i.e., clinically relevant pain, low-intensity clinical pain and clinically irrelevant pain) were not differently distributed (χ² = 1.50, p = 0.826) across visit number, confirming a successful implementation of a pseudorandomized pain state-dependent study design, despite scheduling complexity and pain state transition uncertainty.

**Figure 4.**
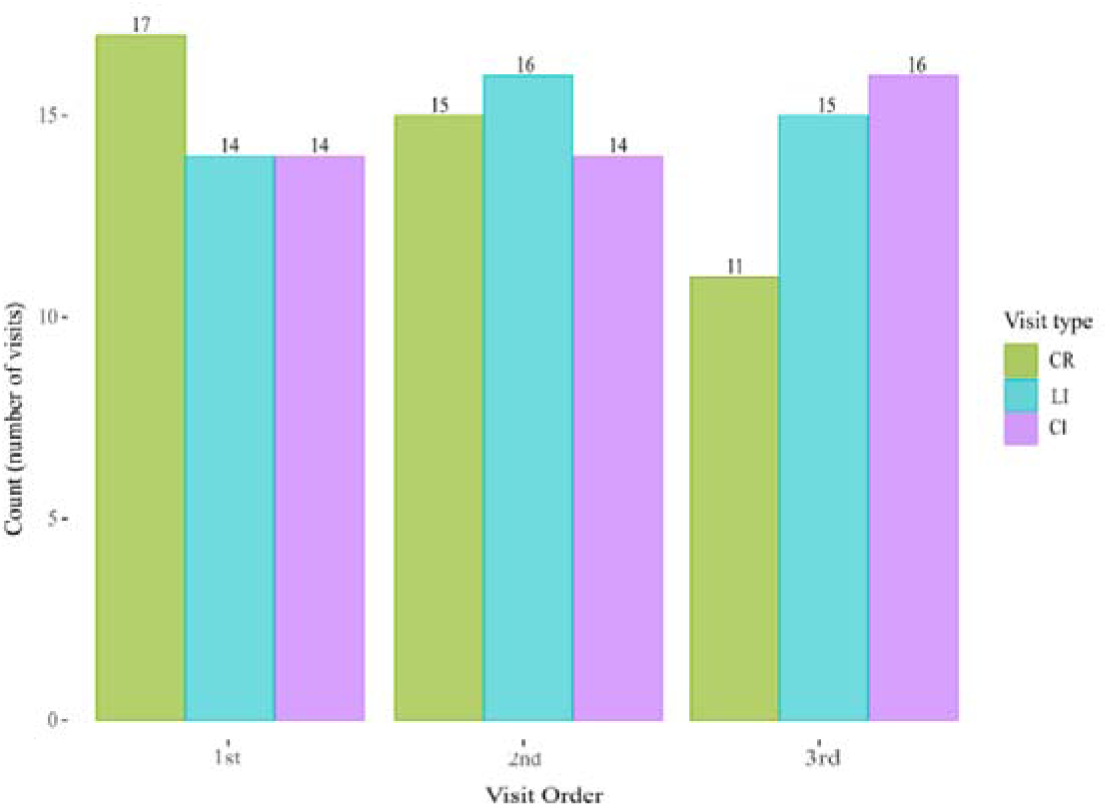
Visit pseudorandomization. Values on top of each bar represent the number of times a certain visit type was carried out in a specific visit order (1^st^, 2^nd^ or 3^rd^). CR: clinically relevant pain, CI: clinically irrelevant pain, LI: low-intensity clinical pain.

At group level, it took approximately two weeks from patient recruitment to the first visit, while the time-interval between visits was approximately three to four weeks (Table 3).

**Table 3.**
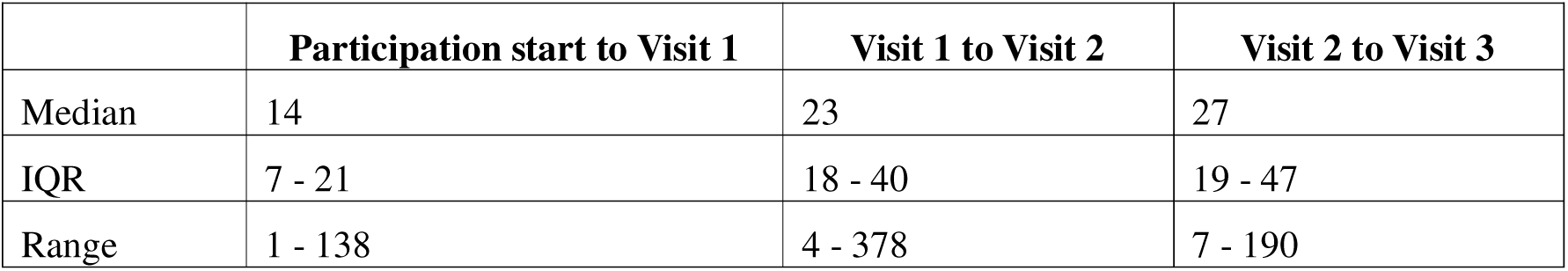
Values are shown in days. Participation start is defined as the initiation of pain intensity tracking with first self-report. IQR: inter-quartile range.

Six mismatches were recorded between the pain state patients were in when arriving on-site and the expected pain state they should have been in for the scheduled visit type. One mismatch happened in the first visit where the patient reported back pain (3.5 NRS) at on-site presentation, as per visit schedule, but gave low pain ratings throughout the whole visit, and most importantly, finished the appointment saying that "actually their back felt really good this day". Therefore, the study team decided a-posteriori to consider this visit as the low-intensity clinical pain one, given that during all testing the pain remained ≤ 3 NRS. The second study visit was then considered the clinically relevant pain visit, as the pain intensity reported at baseline was 5.5 NRS and remained at ≥ 4 NRS throughout the session. Five mismatches happened in the second visit, where at four occasions, patients came in experiencing moderate-to-high back pain when they were expected to have low-intensity back pain, and once vice versa. Because these mismatches happened in the second visit, the scheduled visit type was adapted to match the patient’s actual pain state and the pre-assigned pseudorandomization order was modified a-posteriori to match the new visit sequence.

To further assess whether the prespecified pain intensity criteria for each visit type defined by the study design were achieved, clinical low back pain intensity ratings (0-10 NRS) given by participants at the beginning of the visit were examined (Figure 5, left panel). Pairwise Wilcoxon-tests with Bonferroni corrected p-values confirmed successful pain state separation based on clinical pain intensity, with significantly higher low back pain intensity in the clinically relevant pain visit (mean [SD]: 3.98 [0.90]), compared with both the low-intensity clinical pain (1.03 [0.86]) and the clinically irrelevant pain (1.13 [0.82]) visits (both p-values < 0.001), and no difference between the latter (p = 1.00), as expected. As aimed by participant recruitment, HCs were generally pain-free (0.04 [0.21]), with the exception of two HCs who reported a pain intensity of 1/10 NRS at baseline.

**Figure 5.**
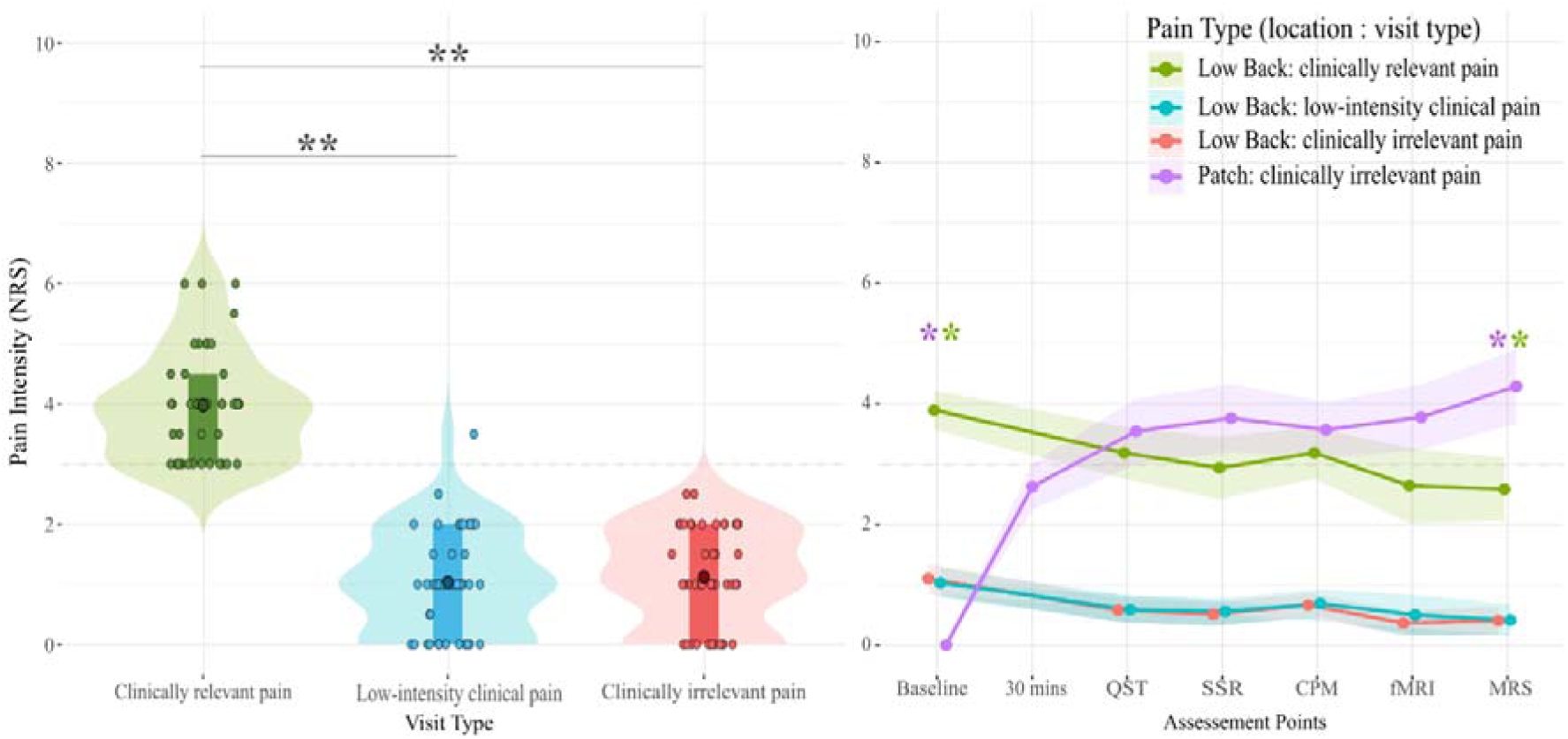
Pain intensity ratings on a 0-10 NRS. The dashed horizontal line (y = 3) represents the threshold delimiting a low back pain episode versus a low-intensity clinical pain / pain free period. Statistical significance is shown with ** (p < 0.001). **Left panel**: individual low back pain intensity ratings reported on-site at baseline. As expected per study design, baseline low back pain intensity ratings were higher in the clinically relevant pain visit than in the low-intensity clinical pain and clinically irrelevant pain visits. **Right panel**: mean (SD) pain intensity across assessment points within a study visit. The assessment points (x-axis) represent the instances at which pain ratings were obtained, i.e., baseline, 30 mins (exclusively for the clinically irrelevant pain visit) and during each of the pain-related measures. Assessment points are shown as discrete occasions reflecting the planned testing sequence rather than as a continuous timeline, as the order of some assessments could vary depending on equipment availability, while the duration of assessments could vary due to individual factors such as differences in understanding instructions or preparation time before each measure. Pain intensity ratings were comparable between the clinically relevant pain state and the clinically irrelevant pain state at all points except at baseline, as expected because the patch had not been applied yet, and during MRS after several hours of exposure to capsaicin. CPM: conditioned pain modulation, fMRI: functional magnetic resonance imaging, MRS: magnetic resonance spectroscopy, NRS: numerical rating scale, SSRs: sympathetic skin responses, QST: quantitative sensory testing.

As a final manipulation check, the induction of clinically irrelevant pain using a Qutenza 8% capsaicin patch as an experimental pain model was investigated. At the beginning of the clinically irrelevant pain visit, patients arrived on-site with low-intensity or no low back pain and with no pain on the dominant arm (0 ± 0 NRS) (Figure 5, right panel). The patch- induced pain started ramping up quickly after application and reached a moderate-to-high intensity during the clinically irrelevant pain visit (Figure 5, right panel).

To identify potential confounding effects of the sheer presence of pain, the goal was to induce experimental pain during the clinically irrelevant pain visit of comparable intensity to the low back pain of the clinically relevant pain visit. This was reached across all measures, except for MRS, where the patch-induced pain was higher than the low back pain (Figure 5, right panel & Table 4). This difference was most likely due to the combination of the two factors, time and testing position. The MRS scan was often the last measure of the visit, and the patch-induced pain intensity was generally higher after several hours of skin exposure to capsaicin. Furthermore, during the MRS scan, patients were laying supine, which is a position that often relieves low back pain. Indeed, all measures that were carried out while participants were laying down, either supine (fMRI and MRS) or prone (SSRs), mean low back pain ratings were < 3 NRS, while for measures carried out in a sitting position (CPM) or both sitting and laying (QST), mean low back pain ratings were > 3 NRS (see Table 4).

**Table 4.** Values represent mean and standard deviation in parenthesis for the low back pain intensity during the clinically relevant pain visit and patch-induced intensity during the clinically irrelevant pain visit, on a 0-10 numerical rating scale, across each measure. P-values are Bonferroni-adjusted; * indicates statistical significance at p < 0.05 for the six Wilcoxon pairwise comparisons shown.

| Assessment Point | Clinically relevant pain (Low back) | Clinically irrelevant pain (Patch) | Test statistic W | Test significance p-value |
| --- | --- | --- | --- | --- |
| Baseline | 3.90 (1.08) | 0 (0) | 0 | < 0.001 * |
| 30 minutes | - | 2.63 (1.31) | - | - |
| QST | 3.19 (1.44) | 3.62 (1.86) | 382 | 1.00 |
| SSRs | 2.94 (1.65) | 3.86 (1.95) | 396.5 | 0.078 |
| CPM | 3.19 (1.44) | 3.59 (1.53) | 349.5 | 1.00 |
| rsMRI | 2.64 (1.87) | 3.78 (1.61) | 297 | 0.057 |
| MRS | 2.58 (1.74) | 4.38 (2.00) | 518 | < 0.001 * |

## 4. Discussion

The operational and implementation considerations arising from the present findings and their implications for study design are discussed in the following sections.

### 4.1 Incorporating Pain Fluctuations in Study Designs

Pain fluctuations are intrinsic to chronic pain, in particular for musculoskeletal pain conditions. However, they are rarely considered in pain research [65], where average pain ratings over different recall periods (e.g., the previous seven days or four weeks) are more commonly used [26,36]. Average pain ratings are also amongst the most frequently used pain assessments in clinical practice, despite the challenges healthcare providers encounter when applying and interpreting them [20]. Considering pain fluctuations may provide additional information beyond average pain intensity by capturing temporal patterns of pain [45]. This is why, in the current study, pain fluctuations were incorporated at different levels of the study design.

Firstly, pain fluctuations were tracked from enrollment until study completion via email using automatic REDCap surveys. The current study employed a time-based sampling approach, which is the most commonly used method in ecological momentary assessment [39]. The selected fixed four-day rating schedule proofed to be sufficient for capturing pain fluctuations for successful scheduling of all three study visits in 93.33% of patients, without imposing excessive burden on participants, indicated by the high median adherence of 93.48%, which was not associated with the self-report burden (i.e., number of expected self-reports).

Although unsuccessful visit scheduling or lower pain rating adherence was an issue for only a few patients, these cases offer important considerations for the optimization of future study designs. As presented in the results section, a four-day fixed rating schedule was not sensitive enough to detect a change in pain state in two patients, and one patient failed to provide pain ratings after the first self-report. However, these three patients were successfully scheduled for all three visits because they proactively contacted the experimenter via other communication channels. In the future, researchers should consider maintaining flexible and open communication with participants, as this may help capture changes that a fixed assessment schedule could fail to detect and provide alternatives for participants who experience difficulties adhering to a specific assessment method. Furthermore, although median adherence was high, the data distribution was broad (ranging from 3.74% to 100%). In fact, this data distribution would be better represented by dividing it into two groups. Most patients (n = 41) showed mid-to-high adherence (range 33.96% – 100%) and successfully reported at least one third of expected pain ratings, while a small group of patients (n = 4) showed very low adherence (range 3.74% to 11.1%). Two patients of this small subgroup did not experience any pain fluctuations for months and stopped reporting their pain intensity at some point, informing the experimenter via other channels. For these two patients, who completed only two out of three visits and remained in a "participating status" for the whole duration of data collection, the planned rating frequency during a period of stable pain resulted in repeated ratings providing limited additional information. In future studies, participant burden could be reduced in such cases by temporarily “freezing” the automatic email reminders or by decreasing their frequency, combined with instructions for participants to contact the experimenter in the event of a clear change in pain state. This approach may also help to improve the homogeneity of adherence data distribution. The other two patients’ low adherence was due to a real lack of self-reports. One patient provided only one rating on the automatic self-report system but contacted the experimenter via other channels. The other patient provided only one rating following the expected schedule. Therefore, these two patients likely struggled to adhere to the self-report method or to the off-site participation requirements. However, they both completed all three study visits, highlighting the benefit of maintaining a flexible self-report system and facilitating open communication between patients and experimenters. Future studies could consider app-based ecological momentary assessment approaches, such as that implemented in the BACPAC study framework [6], which may offer a more user-friendly and accessible interface for repeated self-report, while allowing systematic assessments and potentially incorporating communication with the experimenter within the same platform. This may be particularly useful for participants who experience difficulties with off-site participation requirements.

Secondly, visit scheduling was performed according to pain fluctuations and to current clinical pain state. As discussed above, this procedure worked well, although a high level of flexibility was required to accommodate the availabilities of participants, infrastructure and other involved parties. Furthermore, although patients were instructed to promptly communicate any sudden change in pain state on an appointment day, real-world pain fluctuations sometimes led to patients presenting on-site with a pain state that did not match the one required for the visit type they had been scheduled for. Because these mismatches happened only in the first or second visit, the visit type could in all instances be pragmatically adjusted on the spot and the visit randomization order was adapted a-posteriori. It is important to note that such mismatches between on-site pain state vs. planned visit required spontaneous adjustments which increased experimenter workload (i.e., communicating and coordinating involved parties affected by the change such as nurses, wet lab, and biobank staff, scanner rescheduling, or needing to provide additional documents to clinical examiners). This may have led to incomplete documentation of some visit order changes. Future studies would benefit from a priori standardized procedures for recording such events, or from distributing responsibilities across multiple experimenters, in order to obtain a more objective feasibility metric.

Thirdly, clinical pain intensity was not only recorded on-site at baseline as a confirmation for successful visit type scheduling, but also across different parts of the visit, emphasizing the highly fluctuating nature of back pain. As was presented in the results section, within a stable ongoing pain episode or low-intensity clinical pain period, shorter pain fluctuations occur, often associated with certain positions (i.e. laying vs. sitting), movements (walking, standing, etc.), or context (for example, in an experimental room vs. inside a scanner). This variability presents a challenge when attempting to disentangle state-related from trait-related alterations in musculoskeletal pain conditions, particularly in multimodal assessments where patients are required to assume different body positions. However, as these short-term fluctuations reflect the inherent nature of such conditions, research should not aim to eliminate all sources of variability, but instead acknowledge and account for them when interpreting findings.

### 4.2 An Experimental Pain Model Suitable for Pain Patients

To the best of our knowledge, the Qutenza 8% capsaicin patch has been used as a prolonged experimental pain model (i.e., several hours of application) only in studies with HCs, mainly carried out by laboratories belonging to the same research center [1,22,23,27,60,61,66]. The pain intensity observed in the current study is comparable to values reported in the cited literature, adding to the body of evidence supporting the model’s validity and reproducibility. However, pain intensities reported across individuals are highly variable, highlighting the need for further research into the factors underlying individual differences in responses to this experimental pain model.

The fact that the patch had already been used experimentally and is used therapeutically in pain patients [43,44] ensured the ethical acceptability of this pain model in a patient cohort. The Qutenza 8% capsaicin patch enables the induction of moderate-to-high superficial, burning-like pain, different from clinical low back pain both in location and sensory quality, but similar to clinical pain because it is sustained in time, present at rest, and, based on patients’ comments, can fluctuate with posture changes and movements. Therefore, this pain model is suitable to be used in different chronic pain populations and should be considered in future studies investigating clinical pain-specific alterations from general pain-related changes.

## 5. Conclusion

In conclusion, these results demonstrate the feasibility of a pseudorandomized, pain-based scheduling study design.

## Data Availability

All data produced in the present study are available upon reasonable request to the authors

## Acknowledgements

This work was supported by the Clinical Research Priority Program “Pain” (CRPP Pain) of the University of Zurich (UZH). L.S. is affiliated with the Center for Neuroplasticity and Pain (CNAP), inaugurated by the Danish National Research Foundation (DNRF121) and is supported by a Swiss National Science Foundation Postdoc.Mobility Fellowship (P500- 3_235289/1). The authors acknowledge the help of chiropractors and chiropractor underassistants of the Balgrist University Hospital and the Rückenzentrum Wädenswil, a private chiropractic practice, in participant recruitment. The authors acknowledge the Clinical Trials Unit (CTU) nurses performing the blood sampling. The authors acknowledge the Swiss Center for Musculoskeletal Biobanking (SCMB) for coordinating the provision of study materials for the nurses and for storing the processed blood samples. The authors acknowledge the Swiss Center for Musculoskeletal Imaging (SCMI) and the Magnetic Resonance Center at the University Hospital of Psychiatry (MRZ-PUK) for providing imaging infrastructure and technical support. The authors acknowledge Balgrist Campus and thank the participants for their collaboration.

The data that support the findings of this study are available from the corresponding author upon reasonable request.

The authors have no conflicts of interest to declare.

ChatGPT (OpenAI, version 5.5) was used to assist with R programming and code refinement during data analysis, as well as with language editing and improvement of manuscript readability. The authors reviewed and verified all generated content and take full responsibility for the final manuscript.

## Author contributions

This study protocol was designed by B.C.B., M.H., L.S., and P.S. M.W. was involved in the development of the recruitment process in collaboration with the Chiropractic Policlinic of the Balgrist University Hospital, supported the recruitment process and data monitoring, and set up the momentary pain assessment system. B.W. was involved in the development of the clinical examination. M.W., B.W., and A.L. conducted the clinical examinations. N.Z. set up the magnetic resonance spectroscopy sequence and pre-processed the data that was collected with the help of M.H. S.D. was responsible for the protocol decisions regarding blood sampling and processing. J.D. oversaw the blood sample processing workflow, from receipt of samples from the nurses through processing and storage, processed the majority of samples, and trained other team members involved in the process. B.C.B. recruited participants, scheduled study visits, conducted experiments and scans, analyzed the data, and had a primary role in preparing the manuscript. All authors have approved the final version of the manuscript and agree to be accountable for all aspects of the work.

