## Supplementary Material for "Pain state-dependent multimodal assessment in non-specific low back pain: a pseudorandomized study design with implementation insights"

### Supplementary Material 1 – Baseline Questionnaires

#### Demographics

Date of birth__________________________________

Sex:

☐ Man	☐ Woman	☐ Other

What is your marital status?
☐ Single	☐ Long-term partnership/married	☐ Separated/divorced	☐ Widowed

What is the highest level of education you have compulsory schooling completed?

☐ Vocational apprenticeship	☐ Middle School	☐ Higher Technical School

☐ University of applied sciences	☐ University

What is your current work situation? Multiple answers possible.

(If you are currently unable to work, please state your regular employment relationship)

☐ Working full time (80 - 100%)	☐ Working part-time (< 80%)	☐ Student

☐ Houseman / -wife	☐ Retired	☐ Unemployed	☐ IV - pension

If you are employed: What is your job? __________________________________

What is your mother tongue? Multiple answers possible.

☐ German	☐ French	☐ Italian	☐ English	☐ Spanish	☐ Other. Which?__

How do you rate your German knowledge?

☐ None	☐ Basic knowledge(A1)	☐ Good(B1)	☐ Very good(C1)	☐Fluent(C2)

Have you been unable to work/on sick leave in the last three months due to your back problems?

☐ No	☐ Yes. How many working days (Mon-Fri)? __________________________

If your pain is related to an accident, an occupational illness or claims for compensation (e.g. after an operation), in your opinion are all of them relevant legal or insurance law issues (e.g. compensation for pain and suffering)?

☐ Does not apply	☐ No	☐ Yes

Comments __________________________________________

*You have arrived to the end of the first part. Please press "Submit" to continue.*

#### General Health

##### Disability

This questionnaire has been designed to give us information as to how your back pain has affected your ability to manage everyday life. Please answer every section and mark in each section only the ONE option which applies to you at this time. We realize you may consider 2 of the statements in any section may relate to you, but please mark the box which most closely describes your current condition.

Pain intensity

☐	I have no pain at the moment.

☐	The pain is very mild at the moment.

☐	The pain is moderate at the moment.

☐	The pain is fairly severe at the moment.

☐	The pain is very severe at the moment.

☐	The pain is the worst imaginable at the moment.

Personal Care (washing, dressing, etc.)

☐	I can look after myself normally without causing extra pain.

☐	I can look after myself normally but it causes extra pain.

☐	It is painful to look after myself and I am slow and careful.

☐	I need some help but manage most of my personal care I need help every day in most	aspects of self-care.

☐	I do not get dressed, I wash with difficulty and stay in bed.

Lifting

☐	I can lift heavy weights without extra pain.

☐	I can lift heavy weights but it gives extra pain.

☐	Pain prevents me from lifting heavy weights off the floor, but I can manage if they are	conveniently placed e.g. on a table.

☐	Pain prevents me from lifting heavy weights, but I can manage light to medium weights	if they are conveniently positioned.

☐	I can lift very light weights.

☐	I cannot lift or carry anything at all.

Walking

☐	Pain does not prevent me walking any distance.

☐	Pain prevents me from walking more than 2 km.

☐	Pain prevents me from walking more than 1 km.

☐	Pain prevents me from walking more than 500 meters.

☐	I can only walk using a stick or crutches.

☐	I am in bed most of the time.

Sitting

☐	I can sit in any chair as long as I like.

☐	I can only sit in my favorite chair as long as I like.

☐	Pain prevents me from sitting more than 1 hour.

☐	Pain prevents me from sitting more than 30 minutes.

☐	Pain prevents me from sitting more than 10 minutes.

☐	Pain prevents me from sitting at all.

Standing

☐	I can stand as long as I want without extra pain.

☐	I can stand as long as I want but it gives me extra pain.

☐	Pain prevents me from standing for more than 1 hour.

☐	Pain prevents me from standing for more than 30 minutes.

☐	Pain prevents me from standing for more than 10 minutes.

☐	Pain prevents me from standing at all.

Sleeping

☐	My sleep is never disturbed by pain.

☐	My sleep is occasionally disturbed by pain.

☐	Because of pain I have less than 6 hours sleep.

☐	Because of pain I have less than 4 hours sleep.

☐	Because of pain I have less than 2 hours sleep.

☐	Pain prevents me from sleeping at all.

Sexual Life

☐	My sex life is normal and causes no extra pain.

☐	My sex life is normal but causes some extra pain.

☐	My sex life is nearly normal but is very painful.

☐	My sex life is severely restricted by pain.

☐	My sex life is nearly absent because of pain.

☐	Pain prevents any sex life at all.

Social Life

☐	My social life is normal and gives me no extra pain.

☐	My social life is normal but increased the degree of pain.

☐	Pain has no significant effect on my social life apart from limiting my more energetic	interests e.g. sport.

☐	Pain has restricted my social life and I do not go out as often.

☐	Pain has restricted my social life to my home.

☐	I have no social life because of pain.

Travelling

☐	I can travel anywhere without pain.

☐	I can travel anywhere but it gives me extra pain.

☐	Pain is bad but I manage journeys over two hours.

☐	Pain restricts me to journeys of less than one hour.

☐	Pain restricts me to short necessary journeys under 30 minutes.

☐	Pain prevents me from travelling except to receive treatment.

##### General Health

How do you rate your general health?

☐	excellent

☐	very good

☐	good

☐	less than good

☐	bad

The following question refers to your overall quality of sleep for the majority of nights in the past 7 days. Please think about the quality of your sleep overall, such as how many hours of sleep you got, how easily you fell asleep, how often you woke up during the night (except to go to the bathroom), how often you woke up earlier than you had to in the morning, and how refreshing your sleep was. During the past 7 days, how would you rate your sleep quality overall? (0 = Terrible; 1-3 = Poor; 4-6 = Fair; 7-9 = Good; 10 = Excellent)

☐	0

☐	1

☐	2

☐	3

☐	4

☐	5

☐	6

☐	7

☐	8

☐	9

☐	10

How do you rate the following sentence? It is really not advisable for a person in my condition to be physically active.

☐ Not true	☐ True

##### Substance Consumption

Do you smoke?	☐ No	☐ Yes	☐ Not any more

How many years do you smoke / have you smoked? _____________________________

How many cigarettes (or E-Cigarettes) do you smoke / have you smoked per day? _______

How often have you typically drunk one of the following substances in the last six months?

|  | **Every Day** | **5–6 Days/Week** | **3–4 Days/Week** | **1–2 Days/Week** | **≤3 Times/Month** | **Not at All** |
| --- | --- | --- | --- | --- | --- | --- |
| Wine | ☐ | ☐ | ☐ | ☐ | ☐ | ☐ |
| Beer | ☐ | ☐ | ☐ | ☐ | ☐ | ☐ |
| Liquor | ☐ | ☐ | ☐ | ☐ | ☐ | ☐ |

On a drinking day, what is the typical amount of alcohol consumed?

| Wine | Number of glasses (150 mL): __________ |
| --- | --- |
| Beer | Number of medium beers (33 cl): __________ |
|  | Number of large beers (50 cl): __________ |
| Liquor | Number of shots/alcoholic drinks (44 mL; e.g., vodka, whisky): _________ |

##### Comorbidities

For the disease groups below, please indicate whether you suffer from an illness in the mentioned disease groups and how severely this illness affects you in your everyday life.

|  | **No** | **Yes** |
| --- | --- | --- |
| Heart disease (e.g. angina pectoris, heart attack, cardiac arrhythmia) | ☐ | ☐ |
| Circulatory disease (e.g. high blood pressure) | ☐ | ☐ |
| Lung disease (e.g. asthma, chronic bronchitis) | ☐ | ☐ |
| Gastrointestinal disease (e.g. reflux, stomach ulcer) | ☐ | ☐ |
| Disease of the liver, gall bladder, or pancreas (e.g. hepatitis, gallstones) | ☐ | ☐ |
| Disease of the kidney, urinary tract, or sexual organs (e.g. kidney stones, urinary tract inflammation) | ☐ | ☐ |
| Diseases of the nervous system, brain, or spinal cord (e.g. epilepsy, MS, Parkinson's disease, polyneuropathy) | ☐ | ☐ |
| Metabolic disease (e.g. diabetes) | ☐ | ☐ |
| Skin disease (e.g. psoriasis, neurodermatitis) | ☐ | ☐ |
| Disease of the musculoskeletal system (e.g. rheumatoid arthritis, osteoporosis, osteoarthritis) | ☐ | ☐ |
| Psychiatric disease (e.g. depression, anxiety, schizophrenia) | ☐ | ☐ |
| Tumor/Cancer | ☐ | ☐ |
| Other disease | ☐ | ☐ |

*If yes above:_____________________*

|  | **No Impairment** | **Low Impairment** | **Moderate Impairment** | **Severe Impairment** |
| --- | --- | --- | --- | --- |
| Heart disease | ☐ | ☐ | ☐ | ☐ |
| Circulatory disease | ☐ | ☐ | ☐ | ☐ |
| Lung disease | ☐ | ☐ | ☐ | ☐ |
| Gastrointestinal disease | ☐ | ☐ | ☐ | ☐ |
| Liver, gall bladder, or pancreas disease | ☐ | ☐ | ☐ | ☐ |
| Kidney, urinary tract, or sexual organ disease | ☐ | ☐ | ☐ | ☐ |
| Nervous system, brain, or spinal cord disease | ☐ | ☐ | ☐ | ☐ |
| Metabolic disease | ☐ | ☐ | ☐ | ☐ |
| Skin disease | ☐ | ☐ | ☐ | ☐ |
| Musculoskeletal disease | ☐ | ☐ | ☐ | ☐ |
| Psychiatric disease | ☐ | ☐ | ☐ | ☐ |
| Tumor/Cancer | ☐ | ☐ | ☐ | ☐ |
| Other disease | ☐ | ☐ | ☐ | ☐ |

##### Medication

Do you take medication on a regular basis?	☐ No	☐ Yes

If yes above:

Which medication? __________________________________________

What medication do you take for your back pain? __________________

Comments: __________________________________________

*You have reached the end of this part of the questionnaire. Please press "Submit ".*

#### Pain Characteristics

*The following questions relate to your back pain, even if you have other pain (e.g. neck pain) that is currently the main issue for you. Back pain can lead to back pain and / or pain in the buttocks, leg or foot as well as tingling, numbness or other discomfort in these areas.*

On average, how severe was your pain during the past 4 weeks? Information: If pain has been present for less than 4 weeks, "4 weeks" means since the onset of pain.

☐	0 no pain

☐	1

☐	2

☐	3

☐	4

☐	5

☐	6

☐	7

☐	8

☐	9

☐	10 maximum pain

What do you think is the cause of your pain? ______________________________________

What are your biggest concerns about your pain? __________________________________

##### PainDETECT

Mark the picture that best describes the course of your pain?

☐
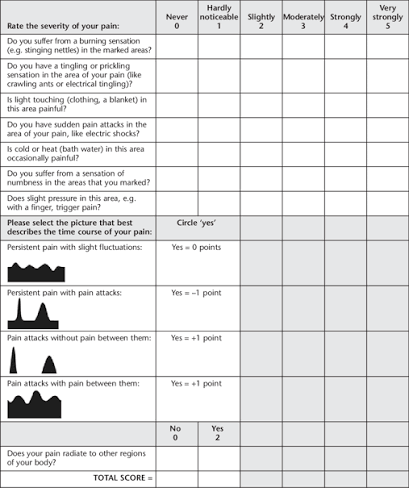
Persistent pain with slight fluctuations

☐
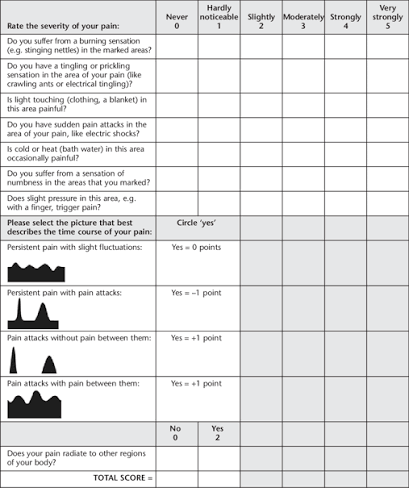
 Persistent pain with pain attacks

☐
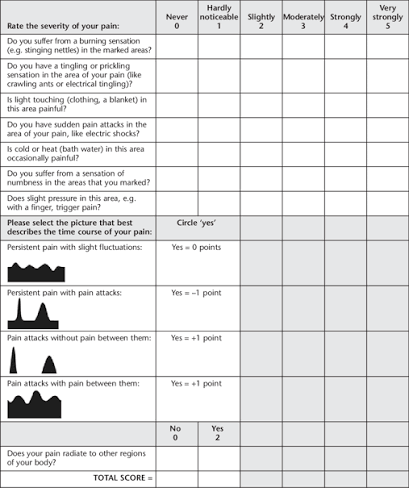
 Pain attacks without pain between them

☐
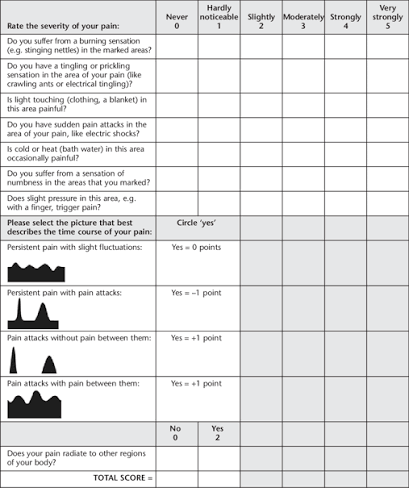
 Pain attacks with pain between them

Does your pain radiate from your main area of pain to other regions of your body?

☐ No	☐ Yes. Where? ________________

In the affected areas of the body (back, leg, buttocks, ...)

|  | **Never** | **Hardly Noticed** | **Slightly** | **Moderately** | **Strongly** | **Very Strongly** |
| --- | --- | --- | --- | --- | --- | --- |
| Do you suffer from a burning sensation (e.g. stinging nettles)? | ☐ | ☐ | ☐ | ☐ | ☐ | ☐ |
| Do you have a tingling or prickling sensation (like crawling ants or electrical tingling)? | ☐ | ☐ | ☐ | ☐ | ☐ | ☐ |
| Is light touching (e.g. clothing, a blanket) painful? | ☐ | ☐ | ☐ | ☐ | ☐ | ☐ |
| Do you have sudden pain attacks like electric shocks? | ☐ | ☐ | ☐ | ☐ | ☐ | ☐ |
| Is cold or heat (e.g. bath water) occasionally painful? | ☐ | ☐ | ☐ | ☐ | ☐ | ☐ |
| Do you suffer from a sensation of numbness? | ☐ | ☐ | ☐ | ☐ | ☐ | ☐ |
| Does slight pressure in this area, e.g. with a finger, trigger pain? | ☐ | ☐ | ☐ | ☐ | ☐ | ☐ |

##### Inflammatory pain pattern

Based on [1]*.* Do the following apply to you?

|  | **No** | **Yes** |
| --- | --- | --- |
| Are your complaints strongest in the morning? | ☐ | ☐ |
| Do you wake up at night because of your symptoms? | ☐ | ☐ |
| Do you feel stiffness for longer than 60 minutes in the morning? | ☐ | ☐ |

##### Widespread Pain Index

Pain in other body regions. Please indicate in which of the listed body regions you have had pain in the last 7 days. Please tick every region of your body in which you had pain, also the region of your back pain.

| ☐ Jaw, left | ☐ Hip (buttock), left |
| --- | --- |
| ☐ Jaw, right | ☐ Hip (buttock), right |
| ☐ Shoulder girdle, left | ☐ Upper leg, left |
| ☐ Shoulder girdle, right | ☐ Upper leg, right |
| ☐ Upper arm, left | ☐ Lower leg, left |
| ☐ Upper arm, right | ☐ Lower leg, right |
| ☐ Lower arm, left | ☐ Neck |
| ☐ Lower arm, right | ☐ Upper back |
| ☐ Chest | ☐ Lower back |
| ☐ Abdomen | ☐ No region |

Indicate the severity of each symptom during the past 7 days.

|  | **No Problem** | **Slight or Mild Problem** | **Moderate Problem** | **Severe Problem** |
| --- | --- | --- | --- | --- |
| Fatigue | ☐ | ☐ | ☐ | ☐ |
| Waking up tired (unrefreshed) | ☐ | ☐ | ☐ | ☐ |
| Trouble thinking or remembering | ☐ | ☐ | ☐ | ☐ |

Other symptoms - in addition to your main pain. Check each of the following other symptoms that you have experienced over the past 7 days:

| ☐ Muscle pain | ☐ Pain in upper abdomen |
| --- | --- |
| ☐ Irritable bowel syndrome | ☐ Nausea |
| ☐ Fatigue | ☐ Nervousness |
| ☐ Thinking or remembering problem | ☐ Chest pain |
| ☐ Muscle weakness | ☐ Blurred vision |
| ☐ Headache | ☐ Fever |
| ☐ Pain / cramps in abdomen | ☐ Diarrhea |
| ☐ Numbness / tingling | ☐ Dry mouth |
| ☐ Dizziness | ☐ Itching |
| ☐ Insomnia | ☐ Wheezing |
| ☐ Depression | ☐ Raynaud (cold hands / cold feet) |
| ☐ Constipation | ☐ Hives / Welts |
| ☐ Ringing in ears | ☐ Vomiting |
| ☐ Heartburn | ☐ Oral ulcers |
| ☐ Loss / change in taste | ☐ Seizures |
| ☐ Dry eyes | ☐ Shortness of breath |
| ☐ Loss of appetite | ☐ Rash |
| ☐ Sun sensitivity | ☐ Hearing difficulties |
| ☐ Easy bruising | ☐ Hair loss |
| ☐ Frequent urination | ☐ Painful urination |
| ☐ Bladder spasms | ☐ None of these symptoms |

##### Central Sensitization Inventory

Please chose the best response for each statement.

|  | **Never** | **Rarely** | **Sometimes** | **Often** | **Always** |
| --- | --- | --- | --- | --- | --- |
| I feel tired and unrefreshed when I wake from sleeping. | ☐ | ☐ | ☐ | ☐ | ☐ |
| My muscles feel stiff and achy. | ☐ | ☐ | ☐ | ☐ | ☐ |
| I have anxiety attacks. | ☐ | ☐ | ☐ | ☐ | ☐ |
| I grind or clench my teeth. | ☐ | ☐ | ☐ | ☐ | ☐ |
| I have problems with diarrhea and/or constipation. | ☐ | ☐ | ☐ | ☐ | ☐ |
| I need help in performing my daily activities. | ☐ | ☐ | ☐ | ☐ | ☐ |
| I am sensitive to bright light. | ☐ | ☐ | ☐ | ☐ | ☐ |
| I get tired very easily when I am physically active. | ☐ | ☐ | ☐ | ☐ | ☐ |
| I feel pain all over my body. | ☐ | ☐ | ☐ | ☐ | ☐ |
| I have headaches. | ☐ | ☐ | ☐ | ☐ | ☐ |
| I feel discomfort in my bladder and/or burning when I urinate. | ☐ | ☐ | ☐ | ☐ | ☐ |
| I do not sleep well. | ☐ | ☐ | ☐ | ☐ | ☐ |
| I have difficulty concentrating. | ☐ | ☐ | ☐ | ☐ | ☐ |
| I have skin problems such as dryness, itchiness, or rashes. | ☐ | ☐ | ☐ | ☐ | ☐ |
| Stress makes my physical symptoms get worse. | ☐ | ☐ | ☐ | ☐ | ☐ |
| I feel sad or depressed. | ☐ | ☐ | ☐ | ☐ | ☐ |
| I have low energy. | ☐ | ☐ | ☐ | ☐ | ☐ |
| I have muscle tension in my neck and shoulders. | ☐ | ☐ | ☐ | ☐ | ☐ |
| I have pain in my jaw. | ☐ | ☐ | ☐ | ☐ | ☐ |
| Certain smells, such as perfumes, make me feel dizzy and nauseated. | ☐ | ☐ | ☐ | ☐ | ☐ |
| I have to urinate frequently. | ☐ | ☐ | ☐ | ☐ | ☐ |
| My legs feel uncomfortable and restless when I am trying to go to sleep at night. | ☐ | ☐ | ☐ | ☐ | ☐ |
| I have difficulty remembering things. | ☐ | ☐ | ☐ | ☐ | ☐ |
| I suffered trauma as a child. | ☐ | ☐ | ☐ | ☐ | ☐ |
| I have pain in my pelvic area. | ☐ | ☐ | ☐ | ☐ | ☐ |

Have you been diagnosed by a doctor with any of the following disorders? Please check the box for each diagnosis and write the year of the diagnosis.

☐	Restless Leg Syndrome

☐	Chronic Fatigue Syndrome

☐	Fibromyalgia

☐	Temporomandibular Joint Disorder (TMD)

☐	Migraine or tension headaches

☐	Irritable Bowel Syndrome

☐	Multiple Chemical Sensitivities

☐	Neck injury (including whiplash)

☐	Anxiety or panic attacks

☐	Depression

☐	None of the above diagnosed by a doctor

Comments __________________________________________

*You have reached the end of this part. Please press "Submit" to continue to the next part.*

### Supplementary Material 2 – Drinking and Smoking Behavior Calculations

#### Drinking behavior

Patients reported the amount of four different alcoholic drinks they consume and the frequency at which they do so. These two variables, are used to calculate the amount of standard drinks / day, based on [2].

Because different types of drinks contain different concentrations of alcohol, the amount of ethanol in each type of drink was calculated:

- Ethanol wine (not weighted for frequency) in mL: amount * 150 (mL standard quantity of glass of wine) * 0.12 (standard 12 % alcohol in table wine)
- Ethanol medium beer (not weighted for frequency) in mL: amount * 330 (mL standard quantity of medium beer) * 0.06 (standard 6 % alcohol in beer)
- Ethanol big beer (not weighted for frequency) in mL: amount * 500 (mL standard quantity of big beer) * 0.06 (standard 6 % alcohol in beer)
- Ethanol liquor (not weighted for frequency) in mL: amount * 44 (mL standard quantity of glass of shot /alcoholic drink) * 0.4 (standard 40 % alcohol in liquors)

Once ethanol quantity for each alcohol type was calculated, this value was multiplied by the weighting value corresponding to consumption frequency of each alcohol type:

- 1: every day
- 0.8: 5-6 days/week
- 0.5: 3-4 days/week
- 0.2: 1-2 days/week
- 0.05: 3 times/month or less
- 0: none at all

This will give the final amount of mL of absolute ethanol / day for each alcohol type. Finally, the sum of all four values of absolute ethanol / day of each alcohol type will give the total ethanol / day consumption.

Because in Switzerland, the amount of mL of ethanol in a standard drink is 10 – 12 mL [3]. To obtain the amount of standard drinks / day the absolute ethanol / day score is divided by 10 (taking the conservative end of the range).

Example: a hypothetical swiss participant who reports drinking 3 glasses of table wine 3-4 days/week, 1 medium beer 1-2 days/week, and 2 mixed drinks each containing a shot of liquor 5-6 days/week. To calculate their average daily ethanol intake (also known as quantity-frequency index, QFI), the frequency weight is multiplied by the quantity and alcohol-based volume for each type of beverage and then add them together. For this hypothetical participant (in mL), we end up with:

Wine: (0.5)(3*150)(0.12) + Beer: (0.2)(33)(0.06) + Liquor: (0.8)(2*44)(0.4) =

27 + 0.396 + 28.16 = 55.56 mL of absolute ethanol/day

Taking 10 mL as the absolute ethanol per drink, a hypothetical swiss participant consumes about 5.5 standard drinks/day.

Based on [5], hazardous drinking behavior was set to drinks / day for women and ≥ 4 drinks / day for men / harmful drinking behavior was set to ≥ 2 drinks / day for women and ≥ 4 drinks / day for men.

#### Smoking behavior

Because one cigarette pack contains 20 cigarettes, pack years are calculated with the following formula:

(cigarettes per day ÷ 20) × years smoked

Based on [4], high risk smoking behavior was set to ≥20 pack-years.
